# Estimation of the number of general anesthesia cases based on a series of nationwide surveys on Twitter during COVID-19 in Japan: A statistical analysis

**DOI:** 10.1101/2020.05.08.20094979

**Authors:** Yosuke Fujii, Hiroki Daijo, Kiichi Hirota

## Abstract

**Purpose:** The coronavirus disease (COVID-19), caused by severe acute respiratory syndrome coronavirus 2 (SARS-CoV-2), has spread to more than 200 other countries. In light of this situation, the Japanese Government declared a state of emergency in seven regions of Japan on April 7, 2020, under the provisions of the law. The medical care delivery system has been under pressure. Although various surgical societies have published guidelines on which to base their surgical decisions, it is not clear how general anesthesia has been performed and will be performed in Japan.

**Methods:** One of the services provided by Twitter is a voting function—Twitter Polls—through which anonymous surveys can be conducted. We used quadratic programming to analyze the results of a series of 17 surveys on Twitter, over the course of 22 weeks. The analysis focused on solving the mathematical optimizing problem on the status of operating restrictions. Public data provided by the Japanese Government were used to estimate the current changes in the number of general anesthesia performed in Japan.

**Results:** The minimum number of general anesthesia per week was estimated at 66.1%, compared to 2015, on April 27, 2020. The time series trend was compatible with the results reported by the Japanese Society of Anesthesiologists.

**Conclusion:** The number of general anesthesia was reduced by up to two-thirds during the COVID-19 pandemic in Japan, and was quantitatively estimated using the Twitter quick questionnaire.

## Introduction

Coronavirus (COVID-19), caused by severe acute respiratory syndrome coronavirus 2 (SARS-CoV-2), was first reported in Wuhan, Hubei, China, and has since spread to more than 200 other countries around the world at the time of writing [1]. In light of this situation, the Japanese Government declared a state of emergency in seven regions of Japan on April 7, 2020, under the provisions of the law. This declaration was extended to the entire nation on April 17, 2020. The incidence of pneumonia from COVID-19 is considerably higher than from seasonal influenza, and the number of cases with no identifiable route of infection has increased rapidly [2, 3]. The medical care delivery system has been under pressure [4]. Emphasis should be placed on maintaining medical care systems. Although various surgical societies have published guidelines on which to base their surgical decisions (https://www.facs.org/covid-19/clinical-guidance/elective-case; https://www.cms.gov/files/document/cms-non-emergent-elective-medicalrecommendations.pdf), it is not clear how general anesthesia has been performed. Social networks such as Twitter are becoming a part of our society, as various information is being accumulated on the Web [5]. One of the services provided is a voting function called Twitter Polls. Using this function, anonymous surveys can be conducted on Twitter in Japan [6].

A series of surveys were conducted on Twitter on the status of operating restrictions. We analyzed the results using quadratic programming to solve a mathematical optimizing problem. Public data provided by the Japanese Government were used to estimate and compare the current changes in the number of general anesthesia carried out in Japan. In addition, the estimated time series trend was validated to be compatible with the results of the surveys sponsored by the Japanese Society of Anesthesiologists (JSA).

## Methods

### Twitter Surveys

Using the Twitter account of @dajhiroki, a board-certified anesthesiologist with approximately 1,300 followers, Twitter Polls was used to conduct seventeen 24-hour surveys, which were held approximately one week apart from each other, from March 13 to August 14, 2020 (Supplementary Information S1). During the period, the spread of COVID-19 had become a social problem in Japan.

The surveys used the same wording throughout the period, in the form of a choice of one of the following four options:

1. No surgical restrictions
2. Partial restrictions (more than half of the usual)
3. Extensive restrictions (less than half of the usual)
4. No scheduled surgery

### Transition of responses to the survey

Suppose there were *R = 4* discrete categories of responses, transition between the responses during the questionnaire period would be estimated (Fig. 1). *P* was a *R × R* transition matrix whose elements *P_ij_* show the probability transition from the *i*th category at time *t* to the *j*th category at time *t + 1*. The transition matrix was

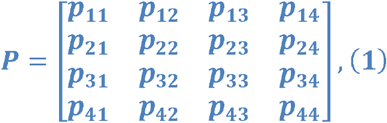

under the constraints of

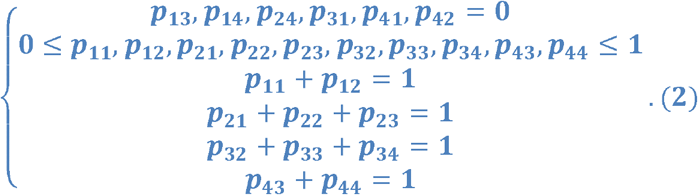

**Figure 1:**
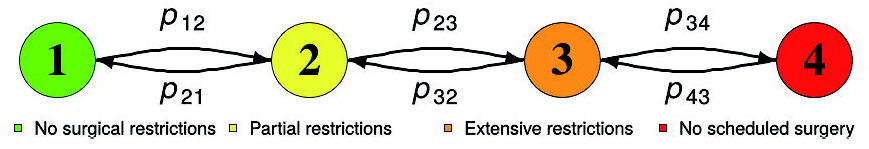
Transition scheme of the proportion of surveys. (1: green) no surgical restrictions, (2: yellow) partial restrictions, (3: orange) extensive restrictions, and (4: red) no scheduled surgery. *p_ij_* is the transition probability from response *i* to response *j*.

The number of the responses to the web questionnaire at time *t* was aggregated as the proportion *Y_i_(t)*, *i = {1,2,3,4}*. When the individual transition was not available, it was not possible to estimate the transition matrix from individual transition data using ordinary least squares method. However, quadratic programming method could estimate the transition matrix from proportional data [7]. The stochastic model of the relation between the actual response counts and the estimated occurrence of *Y_i_ (t)* was described below with the error term *u_j_(t)*;

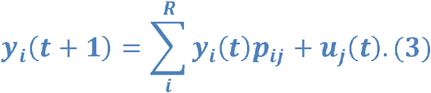

This equation could be written in linear algebraic form as follows:

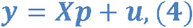

where

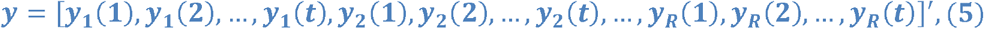

and

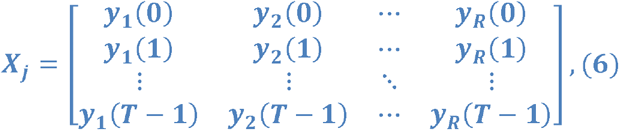

for *j = {1,2,3,4}* so that

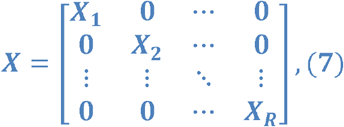

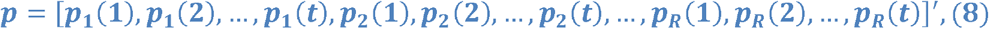

and

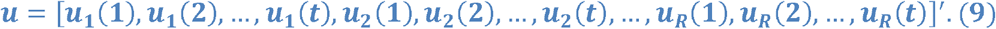

Estimation was performed minimizing the squared error term *|u|^2^* as

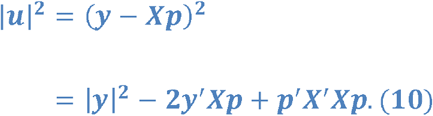

The objective of quadratic programming was to find a vector *p* that would minimize

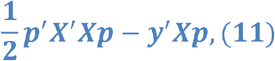

subject to

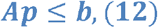

with a real matrix *A* and a real vector *b* as constraints for *p* denoted in equation (2).

### The number of general anesthesia under restriction

The restriction rate of the number of general anesthesia was assumed for each survey response: (1) cases of no surgical restrictions were almost the same as the past statistics, (2) partial restrictions was about 70% restriction rate compared to past statistics, (3) extensive restrictions was about 40% restriction rate compared to past statistics, and (4) no scheduled surgery was about 10% restriction rate compared to the statistics (Fig. 2). Under such assumption, the restriction rate for each hospital *h* at week *t*, *r_t,i,h_* was generated from the probability distribution as follows:

(1) for no surgical restrictive hospitals, *r_t,1,h_* was generated from uniform distribution, *U*, ranging from 0.95 to 1.05.

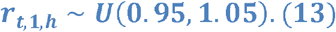

(2–4) for restrictive management hospitals, *r_t,i,h_, i = {2,3,4}*, was generated from beta distribution, *Beta(α,β)*. Their parameters *(α_t,i,h_,β_t,i,h_)*, *i= {2,3,4}* were defined so that the means were 0.7, 0.4, and 0.1, and the variance was 0.005, respectively (Fig. 2).

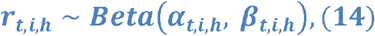

for *i = {2,3,4}*. The number of general anesthesia performed at hospital *h* in the *t*th week, *R_h,t_*, was generated as follows:

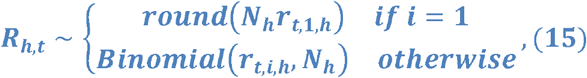

where *N_h_* is the reference number of the general anesthesia performed at each hospital (data available from e-Stat), *round* is a rounding function, and *Bidomial(P,N)* is a binomial sampling function that performs *N* Bernoulli trials with probability *p*.

**Figure 2:**
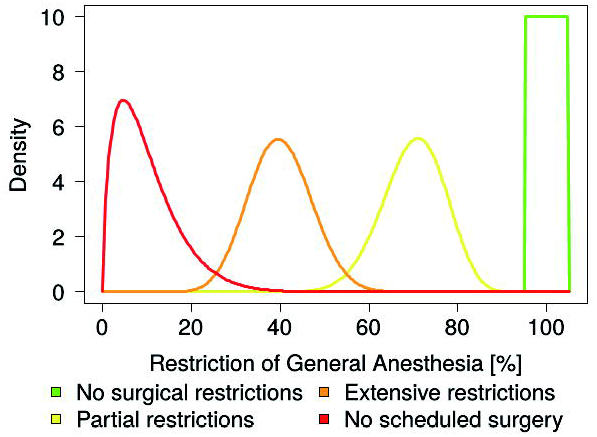
Distribution of the degree of restriction for each response group. The mean percentages of the restrictions were generated from probability distributions. In particular, no surgical restrictions (green) were generated from the uniform distribution *U*(0.95,1.05), while partial restrictions (yellow), extensive restrictions (orange), and no scheduled surgery (red) were generated from the beta distribution, Beta(*α,β*). The set of parameters for beta distribution, (*α,β*), were defined so that the respective means were 0.7, 0.4, and 0.1, and the variances were 0.005.

The survey response for each hospital at *t=1* was randomly allocated to the *i*th response according to *y_i_(t)*, and its status transitioned to the *j*th response by *p_ij_*.

For sensitivity analysis, two other scenarios were considered. An optimistic scenario assumed relatively high performance in operating rooms, with an average of 80% for “2. Partial restrictions,” 50% for “3. Extensive restrictions,” and 25% for “4. No scheduled surgery.” A pessimistic scenario assumed extremely low performance, with an average of 50% for “2. Partial restrictions,” 30% for “3. Extensive restrictions,” and 3% for “4. No scheduled surgery.”

Analysis and estimation were performed using the quadprog package (version 1.5.8) within the R programming language (version 3.4.4). Point estimates and their statistical uncertainty were presented as 95% confidential intervals (CI) with 1000 iteration. The data were obtained from the Japan official statistics portal site e-Stat, a public database in Japan ((https://www.estat.go.jp/stat-search/file-download?statInfId=000031565245&fileKind=0), and were preprocessed for analysis. The code is available online (https://github.com/yfujii08/covid19generalanesthesia) and is available upon request from the corresponding author.

### Nationwide report of the estimation of the number of general anesthesia from the Japanese Society of Anesthesiologists

The JSA determined the extent of the restriction of general anesthesia using questionnaires. They sent questionnaires to all authorized institutes (n=1415) from April 23, 2020, approximately every week. The proportion of the number of general anesthesia compared to 2015 was estimated and disclosed to only the members of JSA (https://anesth.or.jp/img/upload/ckeditor/files/2004_07_08.pdf).

## Ethics

This study is exempted from institutional review board approval because this study does not contain human participants or research material derived from human participants.

## Results

From March 13 to August 14, 2020, responses were solicited from 17 surveys almost weekly via a Twitter account (@dajhiroki) in the Japanese language, with the number of responses received per survey ranging from 47 to 288 (Supplementary Information S1). The results of the surveys are presented in Table 1.

**Table 1:** Results of the Twitter surveys. The sum did not necessarily equal to 100% due to rounding.

| Date of survey (2020) | 3/13 | 3/20 | 3/27 | 4/6 | 4/10 | 4/17 | 4/27 | 5/2 | 5/9 | 5/17 | 5/22 | 6/19 | 7/4 | 7/24 | 7/31 | 8/7 | 8/14 |
| --- | --- | --- | --- | --- | --- | --- | --- | --- | --- | --- | --- | --- | --- | --- | --- | --- | --- |
| Number of votes | 112 | 47 | 73 | 160 | 221 | 232 | 288 | 184 | 93 | 113 | 54 | 157 | 86 | 142 | 100 | 77 | 107 |
| 1. No surgical restrictions | 89% | 77% | 82% | 64% | 46% | 27% | 25% | 29% | 41% | 39% | 46% | 78% | 87% | 80% | 80% | 87% | 82% |
| 2. Partially restricted | 5% | 17% | 7% | 23% | 38% | 38% | 38% | 40% | 37% | 44% | 41% | 19% | 10% | 16% | 11% | 10% | 8% |
| 3. Extensively restricted | 4% | 2% | 4% | 7% | 13% | 27% | 30% | 23% | 19% | 12% | 9% | 2% | 2% | 1% | 4% | 1% | 1% |
| 4. No scheduled surgery | 2% | 4% | 7% | 6% | 3% | 8% | 7% | 8% | 3% | 5% | 4% | 1% | 0% | 4% | 5% | 1% | 8% |

In the public database e-Stat, 3501 hospitals were registered as facilities that provide general anesthesia. Of the 3311 hospitals that reported the number of general anesthesia, 1989 performed more than 100 general anesthesia.

From the survey conducted in the first week (March 13, 2020), 89.2% had no surgical restrictions, but this proportion decreased to 24.7% in the survey conducted in the seventh week (April 27, 2020) then slightly recovered to 38.9% (May 17, 2020). The proportion increased to 87.2% (July 4, 2020) and was finally 80.0% at the end of the questionnaire administration (July 31, 2020). The proportions of partial restrictions, extensive restrictions, and no scheduled surgeries were 5.4%, 3.6%, and 1.8% (March 13, 2020); 37.8%, 29.5%, and 8.0% (April 27, 2020); 44.2%, 11.5%, and 5.3% (May 17, 2020); and 11%, 4%, and 5% (July 31, 2020), respectively (Fig. 3). The estimated proportion of each response and its transition are shown in Fig. 3 and Table 1.

**Figure 3:**
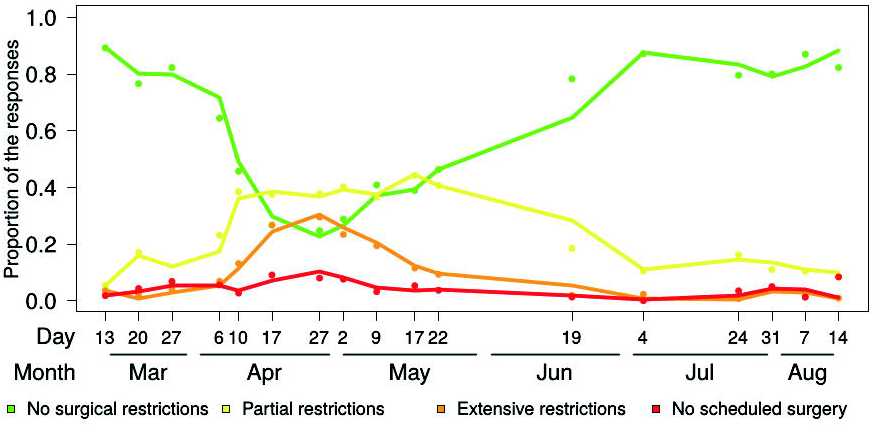
Transition of the proportion of responses to the survey. The solid line was the estimated transition and the dots were actual data.

The number of general anesthesia performed decreased as the restrictions increased (Fig. 4 and Table 2). Before COVID-19, 4.45×10^4^ cases of general anesthesia were performed per week in Japan. The number of general anesthesia (10^4^ per week unit) was estimated to be 4.21 [95% CI: 4.16, 4.26] in the first week (March 13, 2020), 2.95 [2.87, 3.02] in the seventh week (April 27, 2020), and 4.10 [4.04, 4.16] in the last week survey (July 31, 2020) (Table 2). Sensitivity analysis showed that the number of general anesthesia performed decreased according to the intensity of the restrictions (Fig. 5). In the first week survey (March 13, 2020), the optimistic scenario had a median of 4.21×10^4^ /week, while pessimistic scenario had that of 4.14. In the seventh (April 27, 2020) and last week survey (July 31, 2020), the optimistic scenario medians were 3.28 and 4.19, while those of the pessimistic scenario were 2.46 and 3.95, respectively. The maximum difference between optimistic and pessimistic scenarios was 1.34 folds at May 2, 2020.

**Table 2:** Estimated number of general anesthesia (10^4 cases per week) in Japan during the period of the surveys.

| 2020 | 3/13 | 3/20 | 3/27 | 4/6 | 4/10 | 4/17 | 4/27 | 5/2 | 5/9 | 5/17 | 5/22 | 6/19 | 7/4 | 7/24 | 7/31 | 8/7 | 8/14 |
| --- | --- | --- | --- | --- | --- | --- | --- | --- | --- | --- | --- | --- | --- | --- | --- | --- | --- |
| 2.5% | 4.16 | 4.03 | 4.02 | 3.97 | 3.71 | 3.22 | 2.87 | 2.88 | 3.11 | 3.27 | 3.38 | 3.75 | 4.14 | 4.15 | 4.04 | 4.11 | 4.16 |
| 50% | 4.21 | 4.09 | 4.08 | 4.03 | 3.78 | 3.30 | 2.95 | 2.95 | 3.19 | 3.34 | 3.45 | 3.82 | 4.19 | 4.19 | 4.10 | 4.16 | 4.21 |
| 97.5% | 4.26 | 4.14 | 4.14 | 4.08 | 3.84 | 3.36 | 3.02 | 3.03 | 3.26 | 3.42 | 3.52 | 3.88 | 4.23 | 4.24 | 4.16 | 4.21 | 4.25 |

**Figure 4:**
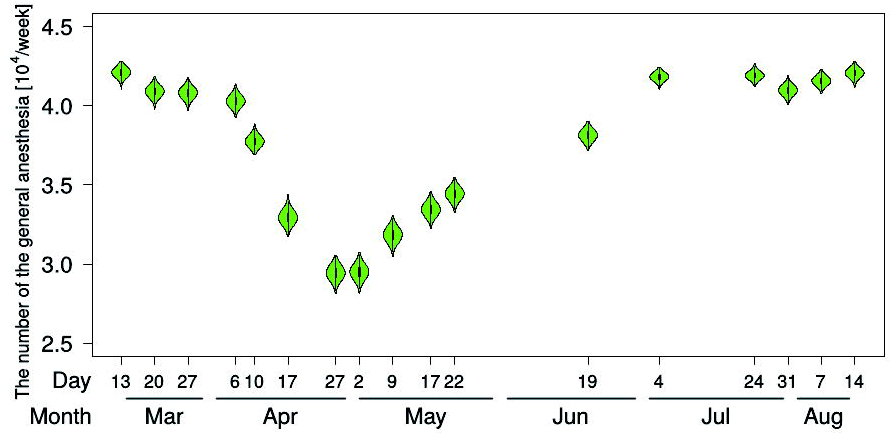
Estimation of the number of general anesthesia in Japan from the survey results. The violinplots show the distribution of the estimated number of general anesthesia performed at 1,989 hospitals in Japan.

**Figure 5:**
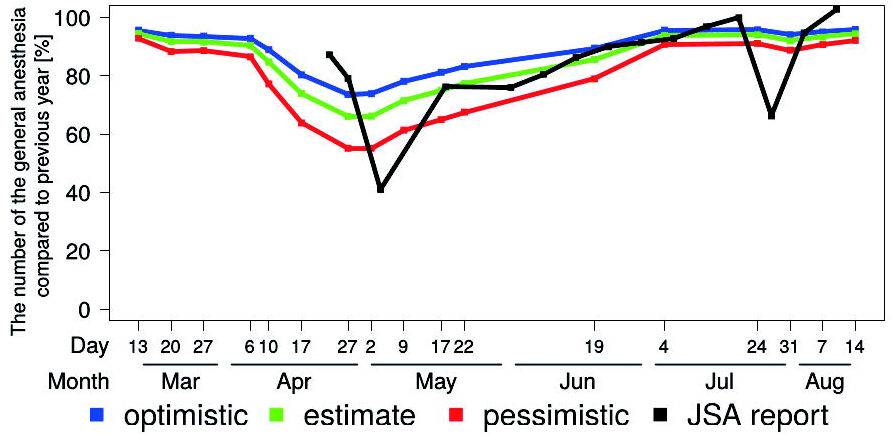
Estimation of the number of general anesthesia in Japan from the results of the surveys for the three scenarios, as a sensitivity analysis and comparison between the estimation of the number of general anesthesia and the report from the Japanese Society of Anesthesiologists (JSA). The JSA reported the results of the estimated number of the general anesthesia from 1415 authorized institutes from April 23, 2020.

The comparison between the estimation of this survey and the nationwide reports from the surveys of the JSA was shown in Fig. 5. The data before the end of April is missing because the JSA questionnaire started from April 23, 2020. The JSA data showed a gradual decrease in the number of general anesthesia compared to 2019: 87.3%, 79.2%, and 41.1% on April 23, April 27, and May 4, respectively. After that, the number of the general anesthesia gradually recovered parallel to the estimation of this survey.

## Discussion

It is difficult to adequately control for the level of limitation of usual surgical care during the spread of COVID-19. It is determined by adherence to official guidelines, restrictions on medical supplies and medical staff, and the level of need for surgery. However, because circumstances can change quickly, it is often determined with reference to the extent of restrictions at a medical facility other than the home facility.

The staged approach—recommended by the Centers for Medicare and Medicaid Services (CMS) and the American College of Surgeons (ACS)—is used as a guide to determine how to perform surgery in situations where the preservation of ventilators and personal protective equipment is necessary, and the ICU has been compromised—or is expected to be compromised soon.

The number of respondents to our surveys increased over time, and it is thought that the number of respondents was dependent on their knowledge of the status of operational limitations. By using a series of surveys conducted through Twitter Polls, we were quickly able to determine the extent of the restrictions on surgery at medical facilities across the country about one month ahead of the survey by the JSA. By periodically soliciting responses to the same survey on Twitter, we were able to estimate the extent to which operations would be restricted nationally—over time.

Our methodology in this study is an estimation method that uses mathematical analysis. We used public data from a database operated by the Japanese Government. The number of hospitals accredited by the JSA is 1415. However, in this study, 1989 hospitals that perform at least 100 surgeries per year under general anesthesia were used. The number of operations performed at each hospital varies—mainly according to the size of the hospital. On the other hand, since neither the size of the hospitals to which the survey respondents belonged nor the changes in the number of operations were known, we used mathematical analysis to calculate the number of operations.

There are several limitations in this study. First, the number and type of survey responses is a potential limitation. It is possible that the number of anesthesiologists using Twitter was not sufficient to obtain a definitive estimation, and there may be some bias in the responses from those that do use it. Responses were low initially, with 47 in the early stages of the surveys, but increased to 288 in the latter half. The growing penetration of the surveys and increasing familiarity with the issue of operating room restrictions may have contributed to an increase in participation, but this may have been biased. Second, we calculated the number of surgeries as 95% to 105% for the criteria: “1. No surgical restrictions,” 70% for “2. Partial restrictions,” 40% for “3. Extensive restrictions,” and 10% for “4. No scheduled surgery.” In the sensitivity analysis, two scenarios were assumed, but the number of general anesthesia performed could be approximately a 1.34-fold difference following the period of the surveys. Because it is difficult to estimate, accurately, the extent of restrictions, intuitive values were adopted. It needs to be compared with the descriptive statistics after the COVID-19 era.

In conclusion, we analyzed the results of a series of surveys on Twitter on the status of operating restrictions using quadratic programming to solve a mathematical optimizing problem. Public data provided by the Japanese Government were used to estimate and compare the current changes in the number of general anesthesia carried out in Japan. The number of general anesthesia was reduced by up to two-thirds during the COVID-19 pandemic in Japan, and was quantitatively estimated using the Twitter quick questionnaire.

## Data Availability

Data is available upon request.

https://github.com/yfujii08/covid19generalanesthesia

## Acknowledgements

We would like to thank Editage (www.editage.com) for English language editing.

## Availability of data and material

All data generated or analyzed during this study are included in this published article. The datasets used and/or analyzed during the current study are available from the corresponding author on reasonable request.

## Ethics declarations

This study is exempted from institutional review board approval because this study does contain no human participants, or research material derived from human participants.

## Additional information

Supplementary Information S1

